# Place Coding in the Human Cochlea

**DOI:** 10.1101/2023.04.13.23288518

**Authors:** Amit Walia, Amanda J. Ortmann, Shannon Lefler, Timothy A. Holden, Sidharth V. Puram, Jacques A. Herzog, Craig A. Buchman

**Author notes:** Corresponding Author: Amit Walia, Washington University School of Medicine, Department of Otolaryngology Head & Neck Surgery, 660 S. Euclid Ave. Campus Box 8115, St. Louis, MO 63110. **Author Contributions:** A.W. designed the study, participated in data collection and analyses, drafted and approved the final version of this paper. C.A.B. designed the study, participated in data collection, drafted and approved the final version of this paper. T.A.H., S.M.L., J.A.H., and A.J.O. participated in data collection, manuscript preparation, and approved the final version of this manuscript. S.V.P. provided critical review of the data and manuscript and approved the final version of the manuscript. **Competing Interest Statement:** C.A.B. serves as a consultant for Advanced Bionics, Cochlear Ltd., Envoy, and IotaMotion and has equity interest in Advanced Cochlear Diagnostics. J.A.H. serves as a consultant for Cochlear Ltd. For the remaining authors, none were declared.

## Abstract

The cochlea’s capacity to decode sound frequencies is enhanced by a unique structural arrangement along its longitudinal axis, a feature termed ‘tonotopy’ or place coding. Auditory hair cells at the cochlea’s base are activated by high-frequency sounds, while those at the apex respond to lower frequencies. Presently, our understanding of tonotopy primarily hinges on electrophysiological, mechanical, and anatomical studies conducted in animals or human cadavers. However, direct *in vivo* measurements of tonotopy in humans have been elusive due to the invasive nature of these procedures. This absence of live human data has posed an obstacle in establishing an accurate tonotopic map for patients, potentially limiting advancements in cochlear implant and hearing enhancement technologies. In this study, we conducted acoustically-evoked intracochlear recordings in 50 human subjects using a longitudinal multi-electrode array. These electrophysiological measures, combined with postoperative imaging to accurately locate the electrode contacts allow us to create the first *in vivo* tonotopic map of the human cochlea. Furthermore, we examined the influences of sound intensity, electrode array presence, and the creation of an artificial third window on the tonotopic map. Our findings reveal a significant disparity between the tonotopic map at daily speech conversational levels and the conventional (i.e., Greenwood) map derived at close-to-threshold levels. Our findings have implications for advancing cochlear implant and hearing augmentation technologies, but also offer novel insights into future investigations into auditory disorders, speech processing, language development, age-related hearing loss, and could potentially inform more effective educational and communication strategies for those with hearing impairments.

**Significance Statement:** The ability to discriminate sound frequencies, or pitch, is vital for communication and facilitated by a unique arrangement of cells along the cochlear spiral (tonotopic place). While earlier studies have provided insight into frequency selectivity based on animal and human cadaver studies, our understanding of the *in vivo* human cochlea remains limited. Our research offers, for the first time, *in vivo* electrophysiological evidence from humans, detailing the tonotopic organization of the human cochlea. We demonstrate that the functional arrangement in humans significantly deviates from the conventional Greenwood function, with the operating point of the *in vivo* tonotopic map showing a basal (or frequency downward) shift. This pivotal finding could have far-reaching implications for the study and treatment of auditory disorders.

## Introduction

Environmental auditory stimuli are complex and encompass a wide range of sound frequencies. The ability to accurately discriminate these frequencies is crucial for effective human communication. Our peripheral auditory organ, the cochlea, features a unique structural layout termed ‘tonotopy’ or place coding, which plays a vital role in frequency discrimination. The auditory hair cells located in the basal (proximal) region of the cochlea, near the round window, are preferentially activated by high-frequency sounds. Conversely, the apical (distal) region’s hair cells exhibit greater sensitivity to lower frequencies. The basilar membrane, along with other soft tissues within the cochlea, acts as a spectral analyzer, spatially separating sound waves based on frequency, leading to distinct points of maximum basilar membrane displacement with resulting hair cell and neural activation. Georg von Békésy was the first to shed light on this spatial specificity by frequency within the cochlea (1-3).

Building on this understanding, when presented with a pure tone stimulus, the basilar membrane undergoes a displacement that peaks at a distinct location before decreasing in amplitude sharply. This displacement results in a unique frequency-to-place map on the basilar membrane, where each cochlear location is optimally responsive to a specific frequency—known as the “best frequency” (BF) or characteristic frequency (CF) when derived at threshold. The path to understanding this tonotopic organization was significantly broadened by the detection of electrical potentials in response to sound, which stem from both the outer hair cells (i.e., cochlear microphonic-CM) and the cochlear nerve’s action potential. These discoveries, demonstrated in cat and guinea pig models, have paved the way for our current understanding of sound transduction (4-8). By integrating von Békésy’s anatomical and physical descriptions with these electrophysiological insights, a fundamental framework has been established for theorizing human sound perception.

Despite these advancements, the electrophysiological characteristics related to frequency discrimination have been largely identified in animal models, severely limiting our understanding of the mechanisms of tonotopy in living humans. The reliability of previous *in vivo* animal and *ex vivo* models have been impeded by several significant and specific challenges. First, the process of surgical alterations and histological processing introduces unavoidable artifacts and spatial discrepancies (1-3, 9, 10). Secondly, the absence of cochlear amplification in *ex vivo* models, which requires the action of outer hair cells to enhance sensitivity and frequency selectivity, hampers their efficacy (11-13). Thirdly, the anatomical and physiological differences between *in vivo* animal models and humans, coupled with the difficulty of deeply probing the cochlear lumen in these models, constrain their applicability (14-16). Lastly, *ex vivo* studies using cadavers do not account for the dynamic biological changes (e.g., cochlear amplifier) that are known to influence passive cochlear mechanics (17). Therefore, *in vivo* electrophysiological measurements within the human cochlea are essential for (1) advancing our knowledge of cochlear tonotopy (2) progressing cochlear implant and hearing augmentation technologies, and (3) improving our understanding of the underlying mechanisms related to auditory disorders.

The primary aim of this research was to elucidate the cochlear tonotopic map in living humans. To address this question, we employed a multi-electrode array, positioned along the longitudinal axis of the cochlear lumen during cochlear implant surgery. This approach has been previously used in cochlear implant patients for assessing hearing preservation (18, 19) and cochlear health and associated speech perception outcomes (20, 21). We initiated the experiment by delivering a pure tone through a sound delivery tube into the ear canal, and then captured the resulting electrophysiologic responses along the multi-electrode array. To comprehend the impact of intensity on the tonotopic map, we modulated the intensity from threshold up to high levels. Subsequently, we evaluated if the insertion of the array within the cochlea induced a perceptual change that would indicate a tonotopic shift. Additionally, we examined if the creation of an artificial ‘third-window’, a procedure frequently employed in e*x vivo* cadaveric and animal experiments, could cause a shift in the tonotopic map. Remarkably, our examination of the *in vivo* tonotopic map in humans revealed that the frequency-position map at conversational sound levels differs significantly from the currently accepted tonotopic map (22-24).

## Results

### *In Vivo* Electrophysiology-Based Frequency-Position Mapping

To construct an *in vivo* electrophysiological map, a multi-electrode array equipped with 22 platinum-iridium electrode contacts was implanted in 50 subjects. Following implantation, computed tomography (CT) imaging was utilized to accurately measure the angular position of each electrode contact, expressed in degrees. The mean number of cochlear turns across the cohort was 2.6, with a range from 2.2 to 2.9 turns. Demographics of all subjects are presented in **Table S1**. Acoustic tone-burst stimuli, ranging between 250 and 4000 Hz and alternating between rarefaction and condensation phases, were introduced immediately post-implantation. This was performed at suprathreshold intensities (∼100 dB sound pressure level [SPL]). Evoked potentials were independently recorded across all 22 electrode contacts. The CM was primarily reflected by the calculated difference between the condensation and rarefaction phases (**Figure 1**). A fast Fourier transformation was applied to these difference waveforms at each electrode, and the amplitude of the first harmonic was evaluated (**Figure 2A**). Using the first harmonic amplitude, CM tuning curves were generated across the electrode array for each subject. The electrode with the largest response on the CM tuning curve was designated the BF. The angular depth of the BF electrodes along the cochlear spiral, as determined by CT imaging, was plotted against the stimulus frequency (**Figure 2B-D**).

**Figure 1.**
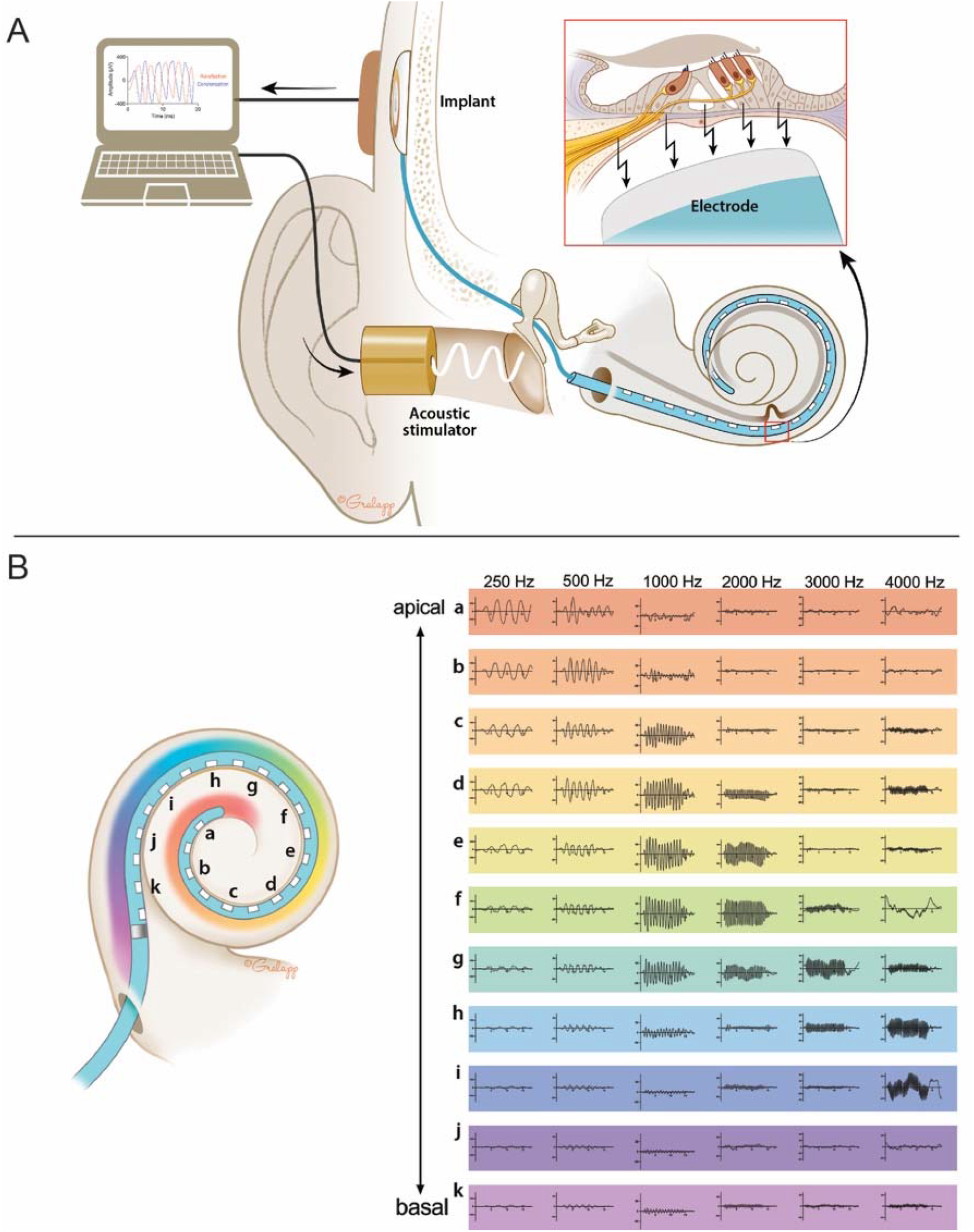
Measurement of live cochlear place coding in humans. (A) An insert earphone was placed into the external auditory canal which was then connected to a sound processor to generate the acoustic stimulus. The surgical site for placement of the recording electrode was then prepared in a sterile-fashion and the round window of the cochlea was exposed using a post-auricular incision and mastoidectomy. The receiver-stimulator for the implant electrode was placed in a subperiosteal pocket and the perimodiolar electrode array was inserted into the round window, within scala tympani according to the manufacturer specification. Following insertion of the electrode, a telemetry coil was placed over the skin in alignment with the cochlear implant antenna which allowed for direct measurement from each of the 22 electrodes on the array in response to acoustically-generated stimulus. The complex signal response measured from each electrode consisted of the electrical activity from outer and inner hair cells and the spiral ganglion (inset). Post-hoc analysis of the ongoing response was performed off-line to separate the hair cell and neural components of the response. A significant response was defined as one whose magnitude exceeded the noise floor by 3 standard deviations. (B) Acoustic stimuli were presented at 250, 500, 1000, 2000, 3000, and 4000 Hz and *in vivo* recordings were made at all 22 electrode contacts. The responses at all even electrodes are shown in the color-matched figure on the right panel where the most-apical electrode (electrode **a**) shows a large response for the 250 Hz stimulus and the responses along more basal electrodes show larger responses for higher frequencies. The electrophysiological properties of the human cochlea are demonstrated with greater electrical responses to higher frequencies at the basal end and larger responses to lower frequencies at the apex. These recordings were performed in 50 subjects, showing similar findings.

**Figure 2.**
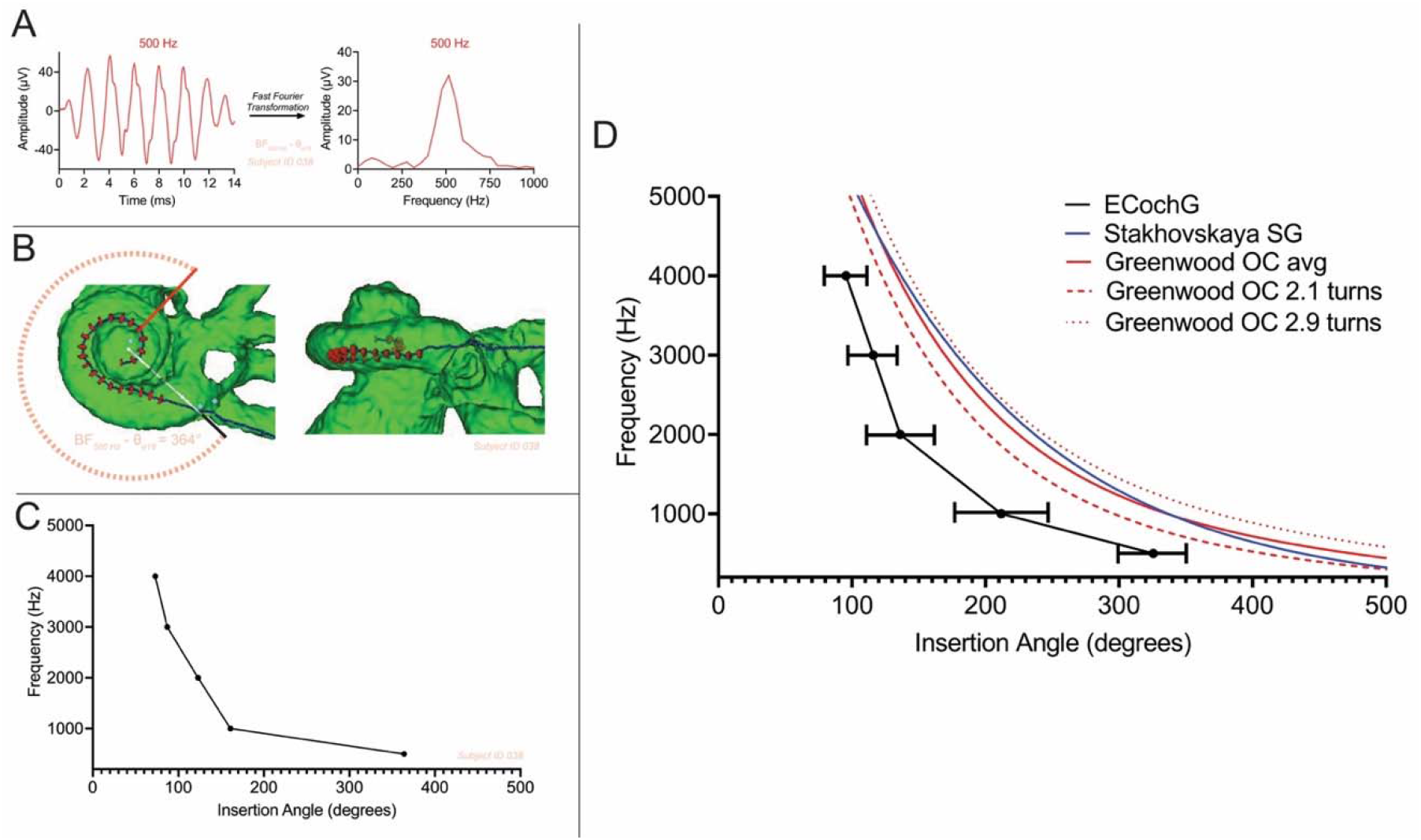
Electrophysiologically-derived frequency-position map. (A) The difference curve was calculated by subtracting rarefaction from condensation phase stimuli. The difference consists primarily of the cochlear microphonic or the ongoing cyclical signal due primarily to the receptor current of outer hair cells. The ongoing portion of the response was selected for fast Fourier transformation and the amplitude of the response to the particular stimulus frequency was determined. Here, 500 Hz is shown for one subject. This was performed across all 22 electrodes in response to 500 Hz and the largest amplitude response was defined as the best frequency (BF) location for that particular frequency (e.g., BF_500 Hz_). In this subject, the BF was at electrode-18 for 500 Hz. (B) Computed tomography (CT) imaging and 3D reconstructions were performed postoperatively to identify the individual electrodes and visualize the adjacent soft tissue anatomy. To determine the position of each electrode, the CT image of each subject’s cochlea was viewed along the mid-modiolar axis and the round window was marked at the 0° at the start of the cochlear canal since all insertions were performed at the round window. The angular position was then measured based on the rotation at the mid-modiolar axis. For this subject, the BF was at electrode-18 which was measured at 364° within the subject’s cochlea. Here, the frequency-position relationship using electrophysiologic responses was determined for 500 Hz. (C) The same methodology described above was performed for 1000, 2000, 3000 and 4000 Hz to develop a frequency-position function for the individual subject’s cochlea using the electrophysiologic responses and CT imaging for the location of each electrode. (D) These electrophysiologic measurements were repeated in 49 additional subjects and CT imaging was performed to identify the precise location of each BF to generate a cumulative frequency-position function for the electrophysiologically-derived map (ECochG map). Error bars are +/-2 standard deviations (SD). This was compared to organ of Corti (OC) and spiral ganglion (SG) maps as established by Greenwood and Stakhovskaya et al., respectively. The ECochG map in this study is at least one octave shifted downward in frequency or more basal in location compared to both the SG and OC maps. The OC map is also shown across various size cochleae (i.e., 2.1 and 2.9 turns), illustrating that the size variability cannot account for the difference between the ECochG map and the OC map.

### Comparison with Preceding Frequency-Position Maps

To assess how the *in vivo* human frequency-position map from the present study deviates from previous models, we compared it with the widely-accepted organ of Corti (OC; Greenwood) and spiral ganglion (SG; Stakhovskaya) maps, (23, 25), (**Figure 2D**). These referenced maps, which form the foundation for understanding cochlear tonotopy, have been recently refined using synchrotron radiation phase-contrast imaging, a technique that allows for enhanced measurements of the cochlea’s helicotrema and hook region (22, 26). Both OC and SG maps preserve frequency separation at levels approximating response thresholds, with minimal divergence in regions where peripheral axons follow a radial trajectory. However, a significant divergence emerges at angles exceeding 600 degrees, a compression point of peripheral axons not reached by the most distal electrode in our study (22).

In examining the five stimulus frequencies (500, 1000, 2000, 3000, and 4000 Hz), a consistent disparity was noted between the BF locations obtained using our *in vivo* map and those estimated by the Greenwood function (**Table S2**). For instance, the Greenwood function predicted the cochlear location for the 500 Hz stimulus to be at 475.6 ± 23.2 degrees, while the *in vivo* measurements revealed an average BF place of 325.6 ± 28.1 degrees (difference 150.0 ± 39.0 degrees). The Greenwood frequency at the 500 Hz BF location was 1083.8 ± 221.4 Hz, resulting in a frequency difference of 583.8 ± 221.4 Hz (1.1 octaves). This frequency and place disparity between *in vivo* recordings and Greenwood tapered basally, reaching 39.4 ± 17.3 degrees and 1666.4 ± 844.5 Hz (0.52 octaves) for the 4000 Hz stimulus.

### Effects of Stimulus Intensity on Frequency Tuning

Previous *in vivo* animal studies have established that increased stimulus sound pressure levels (SPLs) can result in less sharp frequency tuning that shifts the BF position toward a more basal location or where a given location represents a lower BF frequency (27, 28). To further investigate this relationship in humans, we examined the shifts in BF responses across varying stimulus levels in our cohort.

Twenty subjects with moderate to profound residual hearing post-implantation were exposed to varying stimulus intensities, ranging from 36 dB HL to 91 dB HL across a frequency range of 250 Hz to 2000 Hz. Recordings were conducted at the BF electrode and adjacent electrodes to determine if intensity changes would shift the BF location. We found that as the stimulus level increased, the response peak heightened in amplitude but maintained the same location across all frequencies and patients tested. Thus, the frequency tuning and BF location remained stable despite reductions in stimulus intensity, though these responses were limited due to residual hearing, necessitating high stimulus levels (**Figures S1-S3**).

Two subjects, diagnosed with auditory neuropathy spectrum disorder with present distortion product otoacoustic emissions (DPOAEs), represented exceptional cases in which to perform a similar analysis. Auditory neuropathy spectrum disorder is associated with substantial preservation of cochlear hair cell function, thereby avoiding the limitations of our other patients. In both these subjects, increasing the SPL resulted in a broadening peak with the BF location shifting in a basal direction (**Figure 3** and **Figure S4**). Conversely, as the stimulus level was reduced to the equipment’s limits, the expected apical shift, towards Greenwood and Stakhovskaya et al.’s specifications was observed (23, 25). Importantly, when tested with stimulation levels closer to every day conversational speech (∼70 dB HL), responses aligned more closely with those observed during high-level stimulation (i.e. basal shifted) rather than those at threshold. Together, these data suggest the human cochlea’s operating point during typical listening conditions is likely better represented by a map derived from high-intensity stimulation.

**Figure 3.**
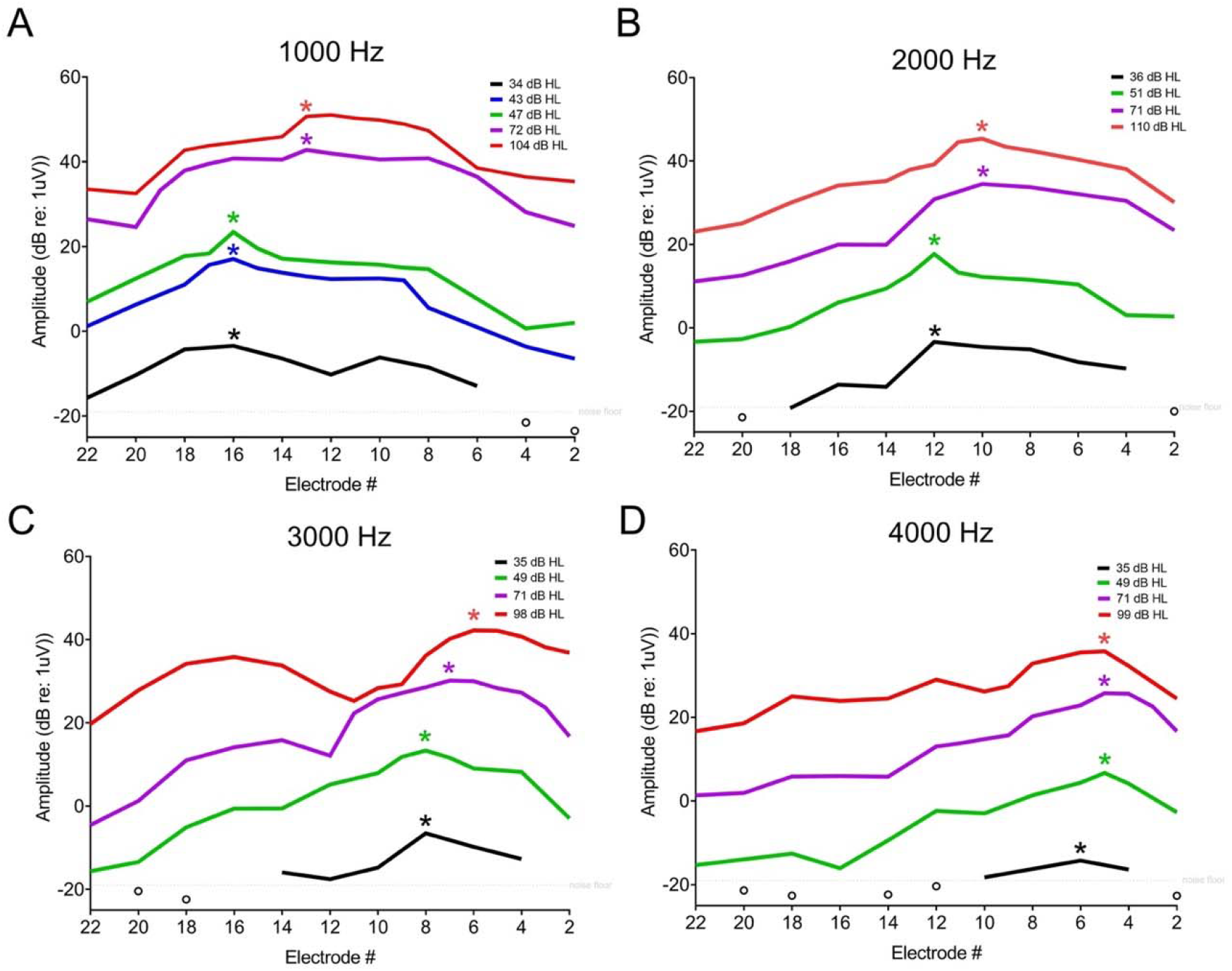
Impact of intensity stimulus on frequency-position map. A subject with auditory neuropathy spectrum disorder was tested, a condition known to have substantial preservation of cochlear hair cell function as evidenced by present distortion-product otoacoustic emissions (2-8 kHz) and cochlear microphonics on auditory brainstem response testing. This was carried out to determine whether stimulus intensity modulation could account for the basal shifted tonotopic tuning derived from the intracochlear electrocochleography (Fig. 2D). Reduction in stimulus level to the limits of the equipment revealed the expected shift in an apical direction for all four frequencies tested: (A) 1000 Hz, (B) 2000 Hz, (C) 3000 Hz, and (D) 4000 Hz. The amplitude shown here are the fast Fourier transformation amplitudes of the difference response, which is primarily representative of the cochlear microphonic tuning curve (i.e., outer hair cell tuning curve). The asterisk (*) represents the best frequency (BF) electrode for each frequency and particular stimulus intensity. The stimulus levels more similar to conversational speech showed responses similar to those seen at highest stimulation level rather than those at threshold, which emphasizes that the frequency-position maps during conversation are more similar to the high-intensity electrophysiologically-derived map rather than that described by the Greenwood equation.

### Impact of Electrode Array on Frequency-Position Map

We proceeded to explore the potential impact of the electrode array on the frequency-position map. Pure tone acoustic stimuli ranging from 125 to 1250 Hz were presented to both ears. To ensure equal loudness across frequencies, we balanced the acoustic stimuli between both ears using a seven-point loudness scale, ranging from inaudible to uncomfortably loud (29). We held one ear constant as the reference, delivering a single pure tone (either 250, 500, or 1000 Hz), while presenting the contralateral ear with pure tones in a random sequence. Subjects were asked to determine whether the pitches presented separately to each ear sounded the ‘same’ or ‘different’ (**Figure S5**). Results showed a minor average difference (range, 1.30-1.65 semitones; 0.11-0.14 octaves) in the acoustic perception of pure tones between both ears in two subjects (**Figure S5B-C**). Therefore, the perimodiolar electrode did not substantially affect the cochlea’s acoustic frequency tuning.

### Effects of Artificial ‘Third-Window’ on Frequency-Position Map

We investigated the potential influence of an artificial cochlear ‘third-window’ on the frequency-position map using an exceptional case. This involved a subject with excellent residual hearing who was scheduled to undergo a translabyrinthine procedure for the resection of a vestibular schwannoma. Prior to labyrinthectomy, the electrode array was inserted into the cochlea’s round window. Acoustically-evoked responses were then measured to determine the BF location across a range of 250 Hz to 4 kHz before and after a fenestration near the upper cochlear turns was created. The creation of the third window did not result in any shift in the frequency-position map (**Figure S6**), suggesting that our recordings were not likely affected by any artifacts from our approach.

## Discussion

### First-ever measurements of human electrophysiologically-derived frequency-position map

This study provides a novel approach to generating an accurate *in vivo* tonotopic map in humans with residual hearing, a task previously unattainable due to the delicate and inaccessible nature of the cochlea. The process of understanding cochlear tonotopy began with von Békésy (1, 2), who meticulously detailed the physical and anatomic observations of the cochlea in response to various tones in human cadavers. This was further advanced by Tasaki et al (4) and Wever et al (7, 8), who pioneered the measurement of electrical potentials from the cochlea in animals, discovering that these potentials were synchronized with the acoustic signal and were a consequence of hair cell stimulation.

Historically, direct measurements of mechanical or neural frequency tuning in cochleae were only feasible in laboratory animals, with assessments of the cochlea’s basilar membrane vibrations largely limited to the basal high-frequency end where surgical access is more convenient (30). Our study leverages cochlear implantation as a unique model for analyzing cochlear mechanics in humans. The strategic placement of the multi-electrode array along the longitudinal axis of the cochlear lumen, in close proximity to residual hair cells and spiral ganglion neurons, enables the collection of robust acoustically-evoked responses. As the number of patients with significant residual acoustic hearing undergoing cochlear implantation increases, so does our ability to obtain substantial responses. This, in turn, allows for a more detailed characterization of the frequency channels established along the entire length of the cochlea, marking a significant advancement in the field of cochlear research.

### *In vivo* map deviates from standard frequency-position functions at conversational intensity levels

The *in vivo* map derived in the present study was subsequently compared to broadly-accepted frequency-position functions, specifically the Greenwood and Stakhovskaya maps (23, 25). Notably, we observed over an octave frequency downward discrepancy (or basal direction shift) between the Greenwood map and the *in vivo* map in the frequency range of 500 to 2000 Hz, with smaller differences at higher frequencies (3 to 4 kHz). Several factors were explored to explain this shift, including stimulus intensity, presence of electrode, and creation of a third window. It is important to note that all subjects in this study had underlying hearing loss, which necessitated their cochlear implants. Consequently, the electrophysiological findings presented here warrant recognition of this clinical context.

In mammalian species, outer hair cells play a crucial role as cochlear amplifiers, enhancing frequency selectivity and auditory sensitivity by up to 40 dB (31, 32). It is reasonably well-documented that high-level stimulation in animals can cause a half-octave shift of the tonotopic map in a basal or frequency-downward direction (33). Additionally, the subjects’ existing otopathology in the present study could potentially impair the active cochlear mechanisms, leading to an additional shift in the tonotopic map (30). These factors likely contribute to our unexpected observation that reductions in stimulus intensity did not shift the tonotopic place coding of the cochlea as expected in most patients, except for the two patients with auditory neuropathy. In these patients, with better preserved amplifier functions, as evidenced by present DPOAEs, the anticipated effect of sound intensity modulation on place coding was observed. Notably, when stimulus intensities were similar to everyday listening conditions (around 70 dB HL), the frequency-position responses aligned more closely with the high-intensity stimulus results, rather than those at threshold. This finding suggests that while our electrophythsiological results would likely align with the Stakhovskaya and Greenwood maps at threshold levels, the map derived from high intensity stimulation is likely more representative of the operating point of the human cochlea during everyday listening conditions.

### Electrode array and artificial third window do not shift frequency-position map

To ensure that the observed basal shift in the frequency-position map in our study was not a result of any intracochlear mechanical impact induced by the electrode array, we carried out inter-aural acoustic pitch comparisons. Although modeled by Kiefer et al (34), previous studies have yet to investigate the possibility of the electrode itself to induce a tonotopic shift in the frequency-position function. If the electrode array itself was causing a shift in the map towards the basal end, we would expect a pitch in the implanted ear to be perceived at a lower frequency than in the ear without an implant. However, our testing of human subjects did not uncover such a shift, suggesting the electrode was not artificially shifting the tonotopic map. . However, it is worth noting that these findings may not apply to lateral wall electrodes, which have a higher likelihood of impacting the basilar membrane (compared to a perimodiolar electrode) due to their limited protection from the osseous spiral lamina (35).

Importantly, we also investigated whether the creation of an artificial ‘third-window’ could potentially introduce an experimental artifact that altered the frequency-position map. While our *in vivo* recordings did not require a third window, von Békésy, in his *ex vivo* observations of the traveling wave, created a fenestration along the bony labyrinth near the cochlear apex to visualize the waveform itself (1-3). Although von Békésy acknowledged the potential for some artifact, particularly at low frequencies, due to this apical fenestration, the impact of the third window has not been thoroughly investigated *in vivo* within the human cochlea (1-3). In an experiment involving a single human subject, we created a third window in a patient with good residual hearing who was undergoing resection of a large vestibular schwannoma via translabyrinthine craniotomy. Interestingly, there was no shift in the frequency-position map with the creation of the third window, suggesting that the observed tonotopic discrepancy could not simply be explained by a third window effect (**Figure S6**). These results also confirmed the robustness of our recordings and the underlying biological basis of our *in vivo* tonotopic map in humans.

### Implications for implanted auditory prosthesesa

While the cochlear implant electrode array was utilized for recordings in this study, its primary function is an auditory prosthesis, designed to electrically stimulate the auditory nerve at a prescribed location. Prior research has underscored the importance of accurate tonotopic stimulation for speech comprehension in complex auditory environments (36, 37). Our study findings are noteworthy since they closely align with those derived in single-sided hearing loss patients using cochlear implants where pitch perception in the normal ear was compared to those from electrically-stimulated contacts from known locations (38, 39).

Recently, discrepancies (or mismatch) between the default frequency allocation algorithms for the individual cochlear implant electrodes and Greenwood’s function have been explored as an explanation for speech perception outcomes in patients (40). This research suggests that although the impact was quite small, greater degrees of mismatch (for frequency-to-place) might negatively influence speech perception outcomes. In our study, we compared the mismatch between our *in vivo* map and the same default frequency allocation tables and discovered a moderate linear correlation where a larger mismatch resulted in poorer cochlear implant speech perception scores (**Figure 4**). When comparing the frequency-to-place mismatch against Greenwood’s frequency-position function in our same patient cohort, we found no correlation with cochlear implant speech perception. These findings bolster the argument that the *in vivo* electrophysiologic map presented in the present study is a far better representation of the actual operating tonotopic map than Greenwood’s function. We believe this to have resulted from Greenwood’s derivation at threshold responses rather than at conversational speech levels. Future research should explore the potential benefits of intensity-based mapping strategies, where different electrodes are activated based on the intensity of the acoustic stimulus, or a strategy that models the tonotopic map close to conversation levels to improve cochlear implant performance. Such strategies could potentially enhance patient outcomes, providing a more effective and personalized approach to cochlear implantation.

**Figure 4.**
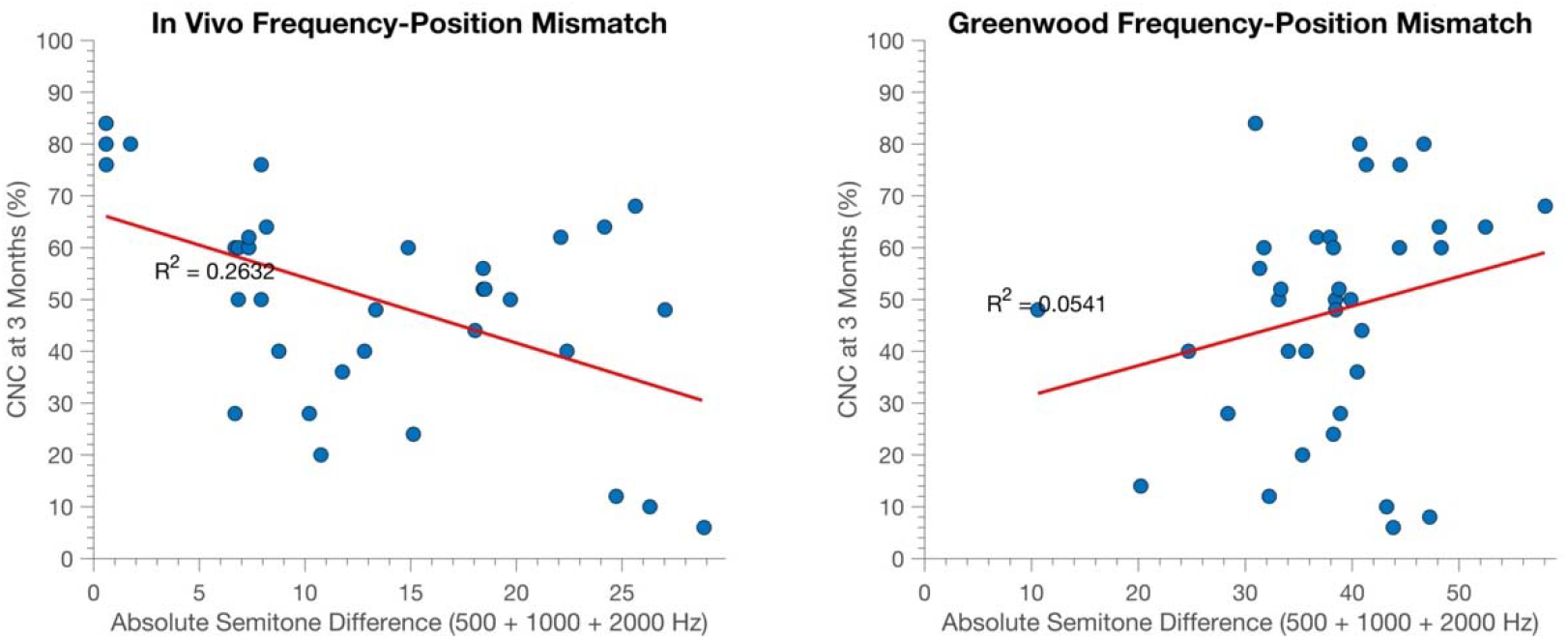
Speech-perception performance following cochlear implantation in relation to frequency-to-place mismatch between *in vivo* and Greenwood maps. We investigated the frequency-to-place mismatch between *in vivo* and Greenwood maps in relation to speech-perception performance post cochlear implantation. The subjects of this study, for whom the *in vivo* map was developed using electrophysiologic recordings and imaging, were evaluated for their cochlear implant performance in a quiet environment at the three months post-activation. The consonant-nucleus-consonant (CNC) word test was employed as an objective performance measure in quiet using the cochlear implant device. In the left panel, we correlate the mismatch (in semitones) between each subject’s default frequency allocation table at the best frequency electrode, as determined by electrophysiologic responses, with CNC word scores. A moderate linear correlation was observed, indicating that lower performance scores were associated with a greater mismatch. In the right panel, we compared the same best frequency electrode with Greenwood’s estimated frequency allocation within the same cohort. We calculated the mismatch from Greenwood’s calculation with the default frequency allocation table provided by the cochlear implant manufacturer. No correlation between these variables was identified. These results reinforce the argument for employing *in vivo* electrophysiologic data for mapping. This methodology may potentially enhance cochlear implant performance compared to the Greenwood map’s place-based mapping approach.

In summary, our findings present the first direct measure and derivation of an *in vivo* tonotopic map in humans. Notably, we discovered that the map, at conversational levels, was shifted by nearly an octave compared to previously established frequency-position maps. This shift is a significant revision to our understanding of the tonotopic map compared with earlier studies. The immediate implication of our findings is for improved mapping and stimulation of cochlear implant electrodes, although there remains broad implications that extend beyond the advancement of cochlear implant and hearing augmentation technologies. Our findings open new possibilities for exploring auditory disorders, speech processing, language development, and age-related hearing loss.

## Materials and Methods

### Study Design and Objectives

This study aimed to (i) construct a frequency-position function based on electrophysiological recordings and compare it with existing organ of Corti and spiral ganglion tonotopic maps of the human cochlea, and (ii) explore how variables such as stimulus intensity, presence of the recording electrode, and the creation of a third window could influence the frequency-position map.

### Participant Selection

Fifty participants were enlisted for this study, with the approval of the Institutional Review Board (IRB) of Washington University in St. Louis (IRB #202007087). Candidates for cochlear implantation were considered as potential participants. Eligibility criteria included adult individuals who possessed residual low-frequency hearing prior to surgery, specifically a low-frequency pure-tone average of 125, 250, and 500 Hz ≤ 60 dB HL. Participants were excluded if they had middle ear pathology, were undergoing revision surgery, or if they lacked a patent external auditory canal, as the acoustic stimulus was delivered via air conduction. Candidates who were not English speaking or were unable to provide informed consent were also excluded.

### Electrode Placement Surgical Procedure

Cochlear implant surgeries were conducted by a team of five experienced surgeons. A standard mastoidectomy-facial recess was used to gain access to the cochlea. Subsequently, the round window niche overhang was partially removed. Depending on the round window membrane orientation, the array was inserted either through a round window incision or after creation of a marginal cochlear opening. All insertions utilized a perimodiolar electrode array (Model CI632; Cochlear Corp., Sydney, NSW, Australia). An intraoperative radiograph confirmed expected coiling of the array. Post-insertion, the cochleostomy was sealed with temporalis muscle or fascia to avert perilymph leakage. The receiver-stimulator was securely positioned in a subperiosteal pocket.

### Intracochlear Electrophysiological Measurements

Prior to the sterilization of the surgical site, an ER3-14A insert earphone (Etymotic, Elk Grove Village, IL, United States) was inserted into the external auditory canal. Once the electrode array was implanted into the cochlea, a telemetry coil was set over the skin, aligned with the cochlear implant antennae using a sterile ultrasound drape. All 22 electrodes within the array were conditioned in reference to the case ground to establish a common reference potential, minimize electrical noise interference, and ensure accurate and reliable measurements in the experimental setup. Tone burst stimuli at frequencies of 250, 500, 1000, 2000, 3000, and 4000 Hz were independently administered in both condensation and rarefaction phases, with a minimum of 30 repetitions per phase. The intensities for the respective frequencies were set at 108, 99.5, 98, 104, 102, and 101 dB HL, determined by the maximum output capacity of the speaker. Each stimulus had a duration of 14 ms with a rise and fall time of 1 ms, shaped by a Blackman window. The recording epoch was set to 18 ms, initiated 1 ms prior to stimulus onset, with a sampling rate of 20 kHz. The electrophysiological responses were recorded across all 22 electrodes of the array.

### Electrophysiological Signal Analysis

The recorded electrophysiological responses, stored as separate condensation and rarefaction phases, were processed offline. Using custom software procedures in MATLAB R2020a (MathWorks Corp., Natick, MA, United States), we calculated the difference curve by subtracting the rarefaction phase stimuli from the condensation phase stimuli. From this difference curve, we selected the ongoing portion of the response for fast Fourier transformation (FFT). This process allowed us to determine the amplitude of the response to the various stimulus frequencies. Utilizing these amplitudes, we were able to generate cochlear microphonic (CM) tuning curves for each frequency across the entire electrode array. In line with previous studies, we defined a significant response as one where the magnitude exceeded the noise floor by three standard deviations (21, 41, 42). The noise floor for this series of recordings was approximately 0.3 μV.

### Determination of Electrode Position using Computed Tomography Imaging

The position of all 22 electrodes along the implanted array was established through post-operative computed tomography (CT) scans and subsequent 3D reconstructions. The platinum iridium contacts cause a “bloom” effect on the CT image, complicating the identification of individual electrodes and adjacent soft tissue anatomy. To overcome this artifact, we employed a validated technique for accurately pinpointing the position of the implanted electrodes within the cochlea (43-45). After co-registering each subject’s pre-implant CT image with their post-implant CT image, electrode contacts were identified and segmented from the post-implant image data and copied onto the pre-implant image space. This composite image was used for subsequent analysis.

To visualize the scalar position of the electrode array and individual electrode contacts, we aligned the composite CT volume with a high-resolution micro-CT cochlear atlas, derived from cadaveric temporal bones (44). This reference was used to infer the location of soft tissue structures within the cochlea that are not resolved by conventional CT (e.g., basilar membrane). The composite CT volume of each subject’s cochlea was viewed along the mid-modiolar axis to determine the position of each electrode. We designated the round window as the 0° starting point of the cochlear canal, since all electrode insertions were performed using a round window-related approach. From this start point, we measured the angular position of each electrode based on rotation about the mid-modiolar axis.

### Electrophysiologically-Derived Frequency-Position Map

The “best frequency” (BF) refers to the specific electrode along the array - corresponding to a particular location on the basilar membrane -that yields the maximum response to a given frequency stimulus. We used the CT-derived angular mapping of the BF electrode for each stimulus to construct a tonotopic map in all 50 subjects. This electrophysiologically-derived map was then directly compared with the established organ of Corti and spiral ganglion frequency-position map functions (22-24).

For a more detailed analysis, the electrophysiology-based map was directly compared to the Greenwood function, with each individual subject’s specific cochlear size taken into account. Our approach began with evaluating the angular location where the Greenwood function predicted the given frequency (250, 500, 1000, 2000, 3000, and 4000 Hz) to be located. Following this, we evaluated the frequency according to Greenwood function at the determined BF location by the electrophysiologically-derived map for the previously defined frequencies. Finally, we computed the discrepancy between the actual and estimated location and frequency, incorporating the octave difference into our calculation.

Additional experiments were conducted to examine the impact of stimulus intensity on the frequency-position map, to investigate pitch discrimination with the electrode array, and to assess the potential impact of a third window on the frequency-position map. These analyses involved a subset of the subjects. A more detailed account of these materials and methods appear in the SI Appendix, Materials and Methods section.

## Data Availability

All data produced in the present study are available upon reasonable request to the authors.

## Acknowledgments

This work was supported by NIH/NIDCD T32DC000022 (A.W.); NIH/NIDCD R01 DC020936 (C.A.B.); American Neurotology Society Research Grant (A.W.).

## Notes

### Funding Statement

AW was supported by NIH/NIDCD institutional training grant T32DC000022. CAB is supported by Department of Defense grant W81XWH19C0054 and NIH/NIDCD grant DC020936-01.

### Author Declarations

Institutional Review Board of Washington University in St. Louis gave ethical approval of this work.

### Summary of Updates

updated figures and main text for clarity

## References

1. G. J. N. L. Békésy, December, Concerning the pleasures of observing, and the mechanics of the inner ear. 11 (1961).

2. G. Von Békésy, Zur Theorie des Hörens: Die Schwingungsform der Basilarmembran (Éditeur inconnu, 1928).

3. G. von Békésy, W. T. Peake (1990) Experiments in hearing. (Acoustical Society of America).

4. I. Tasaki, H. Davis, J. P. Legouix, The Space-Time Pattern of the Cochlear Microphonics (Guinea Pig), as Recorded by Differential Electrodes. The Journal of the Acoustical Society of America 24, 502–519 (1952).

5. H. Davis, The Electrical Phenomena of the Cochlea and the Auditory Nerve. The Journal of the Acoustical Society of America 6, 196–197 (1935).

6. H. Davis, A. J. Derbyshire, M. H. Lurie, L. J. Saul, THE ELECTRIC RESPONSE OF THE COCHLEA. American Journal of Physiology-Legacy Content 107, 311–332 (1934).

7. E. G. Wever, C. W. Bray, AUDITORY NERVE IMPULSES. Science (New York, N.Y.) 71, 215 (1930).

8. E. G. Wever, C. W. Bray, Action Currents in the Auditory Nerve in Response to Acoustical Stimulation. Proceedings of the National Academy of Sciences 16, 344–350 (1930).

9. J. Pichat, J. E. Iglesias, T. Yousry, S. Ourselin, M. Modat, A Survey of Methods for 3D Histology Reconstruction. Medical Image Analysis 46, 73–105 (2018).

10. S. A. Taqi, S. A. Sami, L. B. Sami, S. A. Zaki, A review of artifacts in histopathology. Journal of oral and maxillofacial pathology : JOMFP 22, 279 (2018).

11. D. T. Kemp, Stimulated acoustic emissions from within the human auditory system. The Journal of the Acoustical Society of America 64, 1386–1391 (1978).

12. W. S. Rhode, Some observations on cochlear mechanics. The Journal of the Acoustical Society of America 64, 158–176 (1978).

13. W. E. Brownell, C. R. Bader, D. Bertrand, Y. de Ribaupierre, Evoked mechanical responses of isolated cochlear outer hair cells. Science (New York, N.Y.) 227, 194–196 (1985).

14. P. M. Sellick, G. K. Yates, R. Patuzzi, The influence of Mossbauer source size and position on phase and amplitude measurements of the guinea pig basilar membrane. Hearing Research 10, 101–108 (1983).

15. N. P. Cooper, W. S. Rhode, Basilar membrane mechanics in the hook region of cat and guinea-pig cochleae: Sharp tuning and nonlinearity in the absence of baseline position shifts. Hearing Research 63, 163–190 (1992).

16. J. B. Nadol, Jr., Comparative anatomy of the cochlea and auditory nerve in mammals. Hear Res 34, 253–266 (1988).

17. Y. Zhang et al., Prestin derived OHC surface area reduction underlies age-related rescaling of frequency place coding. Hearing research 423, 108406 (2022).

18. A. Walia et al., Early Hearing Preservation Outcomes Following Cochlear Implantation With New Slim Lateral Wall Electrode Using Electrocochleography. Otol Neurotol 43, 443–451 (2022).

19. A. Walia et al., Is Characteristic Frequency Limiting Real-Time Electrocochleography During Cochlear Implantation? Frontiers in neuroscience 16, 915302 (2022).

20. A. Walia et al., Electrocochleography and cognition are important predictors of speech perception outcomes in noise for cochlear implant recipients. Scientific Reports 12, 3083 (2022).

21. A. Walia et al., Promontory Electrocochleography Recordings to Predict Speech-Perception Performance in Cochlear Implant Recipients. Otol Neurotol 43, 915–923 (2022).

22. L. Helpard et al., An Approach for Individualized Cochlear Frequency Mapping Determined From 3D Synchrotron Radiation Phase-Contrast Imaging. IEEE Trans Biomed Eng 68, 3602–3611 (2021).

23. D. D. Greenwood, A cochlear frequency-position function for several species--29 years later. J Acoust Soc Am 87, 2592–2605 (1990).

24. O. Stakhovskaya, D. Sridhar, B. H. Bonham, P. A. Leake, Frequency map for the human cochlear spiral ganglion: implications for cochlear implants. Journal of the Association for Research in Otolaryngology : JARO 8, 220–233 (2007).

25. O. Stakhovskaya, D. Sridhar, B. H. Bonham, P. A. Leake, Frequency Map for the Human Cochlear Spiral Ganglion: Implications for Cochlear Implants. Journal for the Association for Research in Otolaryngology 8, 220 (2007).

26. H. Li et al., Three-dimensional tonotopic mapping of the human cochlea based on synchrotron radiation phase-contrast imaging. Scientific Reports 11, 4437 (2021).

27. M. Chatterjee, J. J. Zwislocki, Cochlear mechanisms of frequency and intensity coding. I. The place code for pitch. Hear Res 111, 65–75 (1997).

28. I. J. Russell, K. E. Nilsen, The location of the cochlear amplifier: spatial representation of a single tone on the guinea pig basilar membrane. Proc Natl Acad Sci U S A 94, 2660–2664 (1997).

29. P. J. Blamey, L. F. Martin, Loudness and satisfaction ratings for hearing aid users. J Am Acad Audiol 20, 272–282 (2009).

30. L. Robles, M. A. Ruggero, Mechanics of the mammalian cochlea. Physiol Rev 81, 1305–1352 (2001).

31. M. C. Liberman et al., Prestin is required for electromotility of the outer hair cell and for the cochlear amplifier. Nature 419, 300–304 (2002).

32. R. Fettiplace, C. M. Hackney, The sensory and motor roles of auditory hair cells. Nature reviews. Neuroscience 7, 19–29 (2006).

33. M. A. Ruggero, N. C. Rich, A. Recio, The effect of intense acoustic stimulation on basilar-membrane vibrations. Auditory Neuroscience 2, 329–345 (1996).

34. J. Kiefer, F. Böhnke, O. Adunka, W. Arnold, Representation of acoustic signals in the human cochlea in presence of a cochlear implant electrode. Hear Res 221, 36–43 (2006).

35. F. Risi, Considerations and Rationale for Cochlear Implant Electrode Design - Past, Present and Future. The journal of international advanced otology 14, 382–391 (2018).

36. A. J. Oxenham, J. G. Bernstein, H. Penagos, Correct tonotopic representation is necessary for complex pitch perception. Proc Natl Acad Sci U S A 101, 1421–1425 (2004).

37. M. K. Qin, A. J. Oxenham, Effects of simulated cochlear-implant processing on speech reception in fluctuating maskers. J Acoust Soc Am 114, 446–454 (2003).

38. J. P. M. Peters, E. Bennink, G. A. van Zanten, Comparison of Place-versus-Pitch Mismatch between a Perimodiolar and Lateral Wall Cochlear Implant Electrode Array in Patients with Single-Sided Deafness and a Cochlear Implant. Audiology and Neurotology 24, 38–48 (2019).

39. T. F. Tóth et al., Matching the pitch perception of the cochlear implanted ear with the contralateral ear in patients with single-sided deafness: a novel approach. Eur Arch Otorhinolaryngol 10.1007/s00405-023-08002-z (2023).

40. M. W. Canfarotta et al., Frequency-to-Place Mismatch: Characterizing Variability and the Influence on Speech Perception Outcomes in Cochlear Implant Recipients. Ear Hear 41, 1349–1361 (2020).

41. D. C. Fitzpatrick et al., Round window electrocochleography just before cochlear implantation: relationship to word recognition outcomes in adults. Otology & neurotology : official publication of the American Otological Society, American Neurotology Society [and] European Academy of Otology and Neurotology 35, 64–71 (2014).

42. N. H. Calloway et al., Intracochlear electrocochleography during cochlear implantation. Otol Neurotol 35, 1451–1457 (2014).

43. L. K. Holden et al., Factors affecting open-set word recognition in adults with cochlear implants. Ear and hearing 34, 342–360 (2013).

44. J. Teymouri, T. E. Hullar, T. A. Holden, R. A. Chole, Verification of computed tomographic estimates of cochlear implant array position: a micro-CT and histologic analysis. Otol Neurotol 32, 980–986 (2011).

45. M. W. Skinner et al., In vivo estimates of the position of advanced bionics electrode arrays in the human cochlea. The Annals of otology, rhinology & laryngology. Supplement 197, 2–24 (2007).

