## Supplementary File for "Place Coding in the Human Cochlea"

**Materials and Methods**

**Assessing the Impact of Stimulus Intensity on Frequency-Position Map**

The influence of stimulus intensity on cochlear frequency tuning was examined through electrophysiological recordings in 22 subjects, which included twenty from the initial pool of 50 subjects and two with auditory neuropathy spectrum disorder. After identifying the BF electrode at the highest intensity stimulus (determined by the speaker's limit) for a specific frequency, we reduced the stimulus intensity in 5-dB increments. We conducted these measurements at the BF electrode and the adjacent electrodes to ascertain whether a decrease in stimulus intensity would lead to an apical or basal shift of the BF. For these measurements, a single sweep was performed and the noise floor was set at ~1.0 μV. All other parameters related to stimulus and recording, including signal processing, were identical to the procedure described earlier.

In order to model the effects of stimulus intensity changes on BF place in a cochlea with significantly preserved hair cell function, we selected two patients with auditory neuropathy spectrum disorder. These patients exhibited both present distortion-product otoacoustic emissions (DPOAEs) between 2 and 8 kHz and auditory brainstem response waveforms characteristic of a CM with absent neural waveforms. Post electrode array placement, measurements were taken across every electrode along the array at high-intensity (~90 dB HL, conversation-level intensity (~70 dB HL), and near threshold (~20-30 dB HL) for five stimulus frequencies (500-, 1000-, 2000-, 3000, and 4000-Hz). For these subjects, thirty sweeps were performed, and the CM tuning curves were generated for varying intensities and frequencies.

**Examining Pitch Discrimination with Electrode Array**

In order to investigate the electrode's influence on the frequency-position map, we conducted acoustic pitch-discrimination comparisons between ears in two subjects with unilateral cochlear implants. The cochlear implant processor was not utilized during this part of the test. Both subjects had been implanted for less than six months and had preserved residual hearing in the implanted ear after the operation (postoperative low-frequency pure tone average of 125-, 250-, and 500-Hz <60 dB HL). Furthermore, both subjects had comparable residual hearing in the non-implanted ear.

Each participant attended two sessions, with a minimum interval of two weeks between them. Each session lasted approximately two hours and took place in a double-walled sound booth. Audiometric air conduction thresholds ranging from 250 Hz to 8 kHz were recorded using insert earphones for each ear. We first balanced the loudness of the acoustic stimuli at the tested frequency between ears, employing a seven-point loudness scale ranging from inaudible to uncomfortably loud (29). This allowed us to confirm that the stimulus intensity was comfortable for both ears. Subsequently, we kept the stimulus intensity of the non-implanted ear constant at a comfortable level, while adjusting the acoustic stimulus intensity of the implanted ear in 5-dB and then 1-dB increments to pinpoint the exact stimulus level at which the tone sounded similar in both ears. This process was repeated three times for each frequency tested (250-, 500-, 1000-Hz), and we used the median value for the pitch-discrimination part of the test.

After achieving loudness balance at a comfortable level, we kept the tested frequency (250-, 500-, 1000-Hz) constant in the non-implant ear, while presenting the implant ear with acoustic pure tones that spanned 2 octaves from the non-implant ear's constant frequency, in a randomized order. A total of 80 individual presentations of various pure tones were made. The subject was then asked to specify whether the pitches presented to each ear separately sounded 'same' or 'different'. We defined outliers as instances where the subject's response deviated from the other four responses, an occurrence often attributed to fatigue in pitch-discrimination testing (45). This procedure was repeated in a separate session, but this time, the implant ear frequency remained constant while the non-implant ear frequency was varied.

**Assessing the Impact of a Third Window on the Frequency-Position Map**

Previous *in vitro* studies investigating tonotopy, including those conducted by von Békésy, have utilized a fenestration of the cochlear lumen for direct observation or recording of the traveling wave. However, the potential influence of this artifact on the frequency-position map remains unexplored. To assess the effect of a third window on electrophysiologic recordings, we conducted a study involving a human subject with excellent residual hearing. This subject was undergoing a translabyrinthine craniotomy, a procedure that intentionally destroys residual hearing, for the removal of a vestibular schwannoma.

Before the labyrinthine resection, the cochlea was accessed and the same type of device as described above was inserted. The BF was identified along the array for stimulus frequencies of 250-, 500-, 1000-, 2000-, 3000-, and 4000-Hz at the maximum intensity output of the speaker. The signals were processed following the previously described methods. After generating the CM tuning curves, a 1-mm diamond burr was used to create a fenestration near the upper cochlear turns. Subsequently, CM tuning curves were generated again to investigate if the third window had altered the location of the BF. Following these procedures, the implant was removed, and the surgery proceeded without complications. The CM tuning curves were generated for individual frequencies to allow a comparison between pre- and post-fenestration of a third window.

**Supplementary Figures**

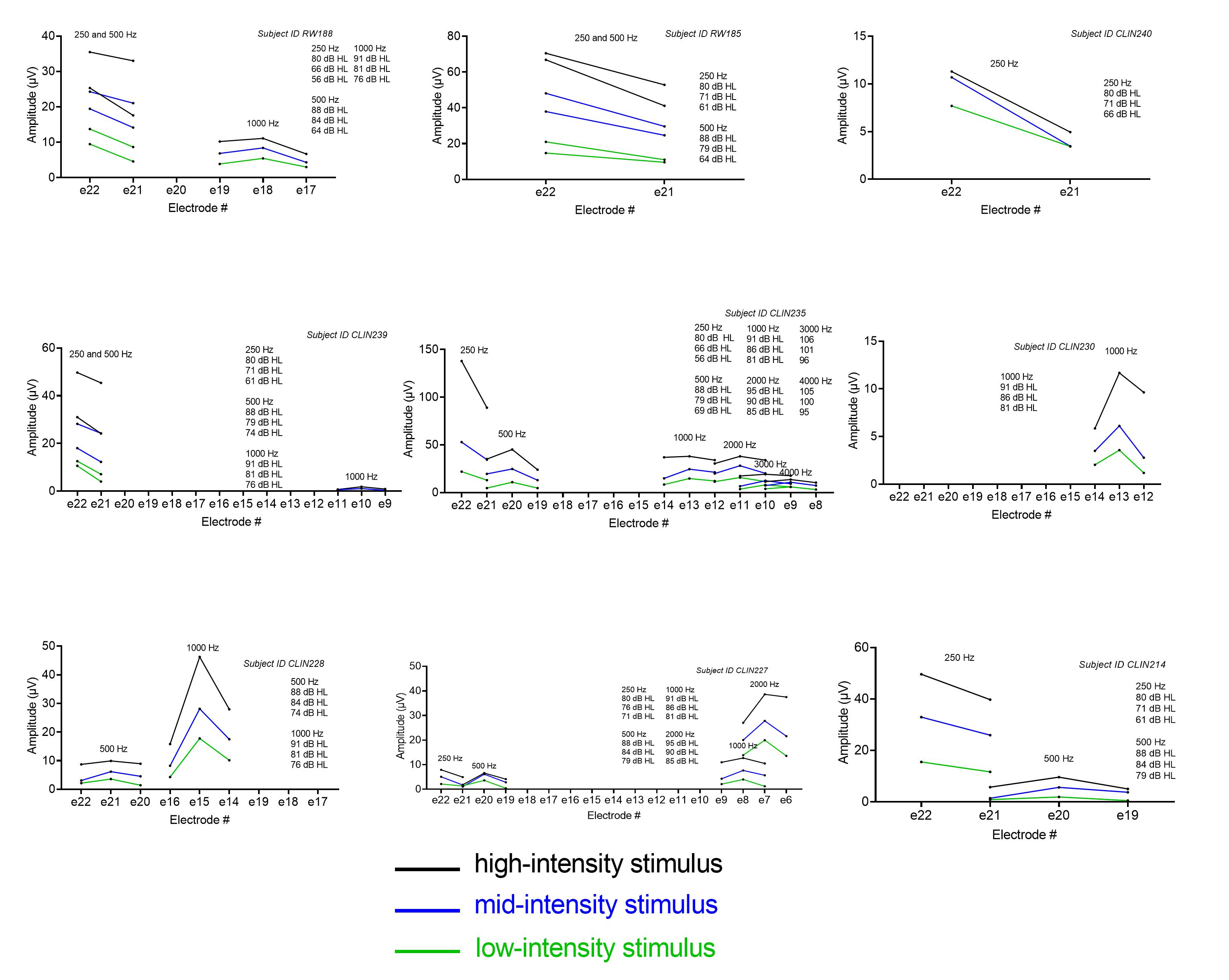

**Fig. S1. Stimulus intensity and the frequency-position map, Part A.** The impact of stimulusintensity on the frequency tuning curve of the cochlea was evaluated in 20 subjects. Once the best frequency (BF) electrode was identified at the highest intensity stimulus (as defined by the limit of the speaker) for a particular frequency, the stimulus intensity was decreased and measurements were performed at the BF electrode and immediately adjacent electrodes to determine whether there would be an apical shift or basal shift of the BF with decreasing stimulus intensity. The frequency tuning and location of the BF did not shift with decreases in stimulus intensity, albeit responses were limited by the amount of residual hearing which necessitated high stimulus levels, in 20 subjects that we tested.

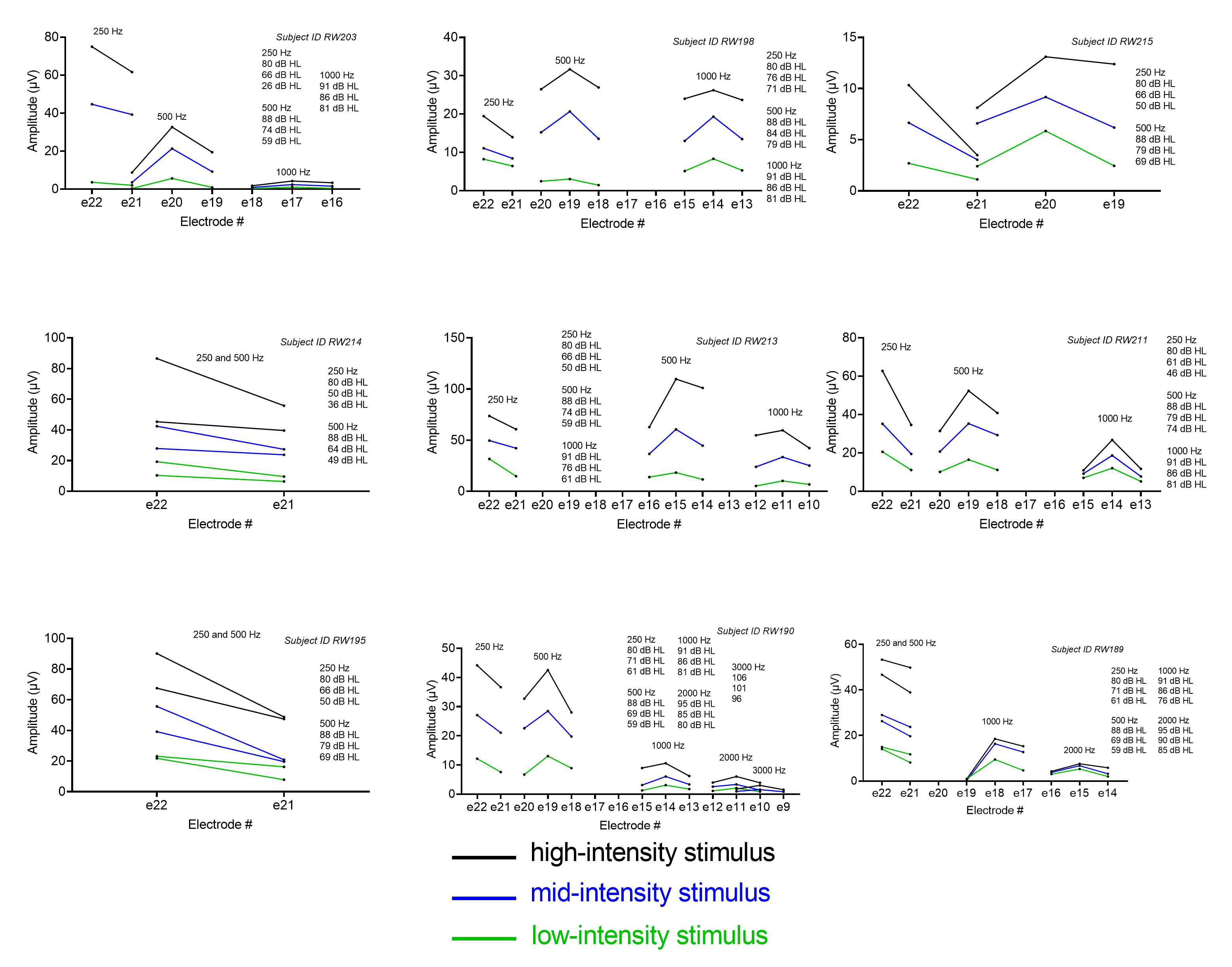

**Fig. S2. Stimulus intensity and the frequency-position map, Part B**

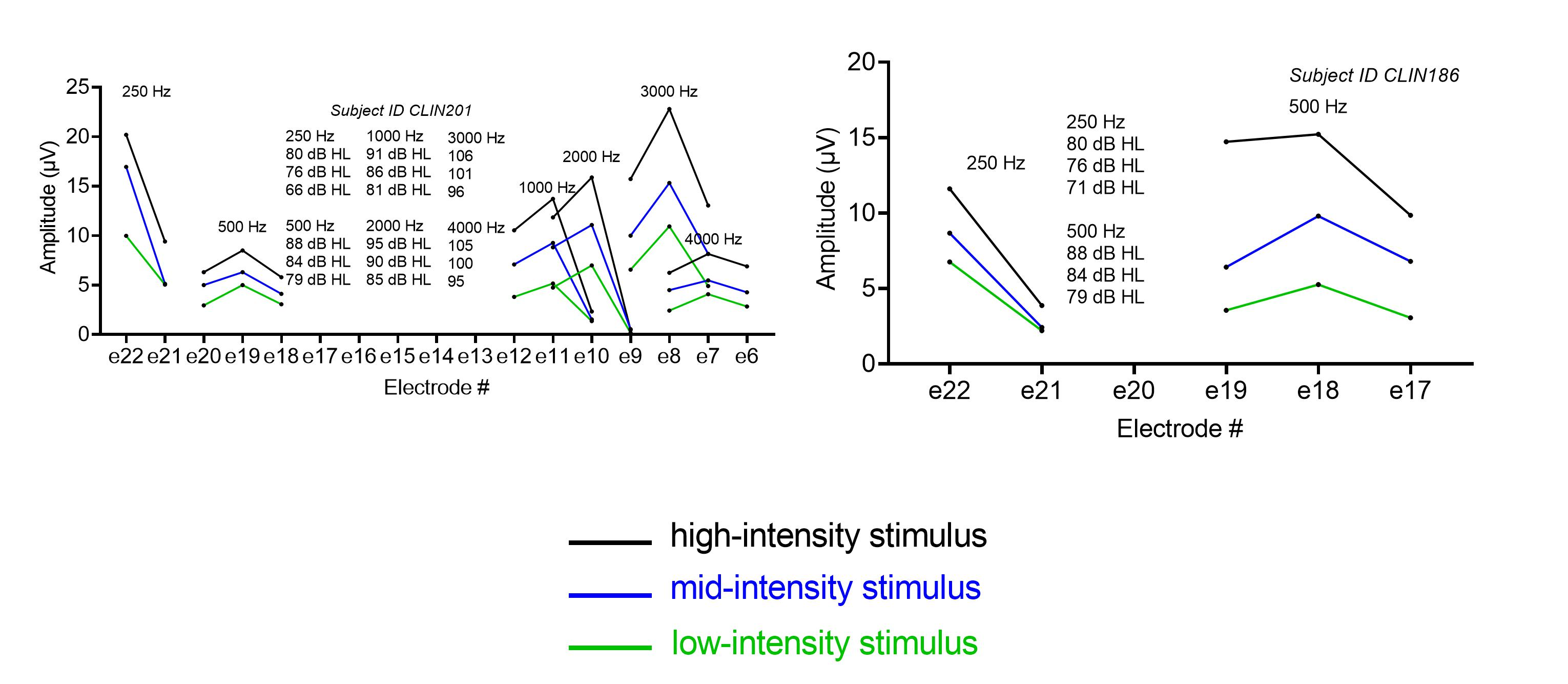

**Fig. S3. Stimulus intensity and the frequency-position map, Part C.**

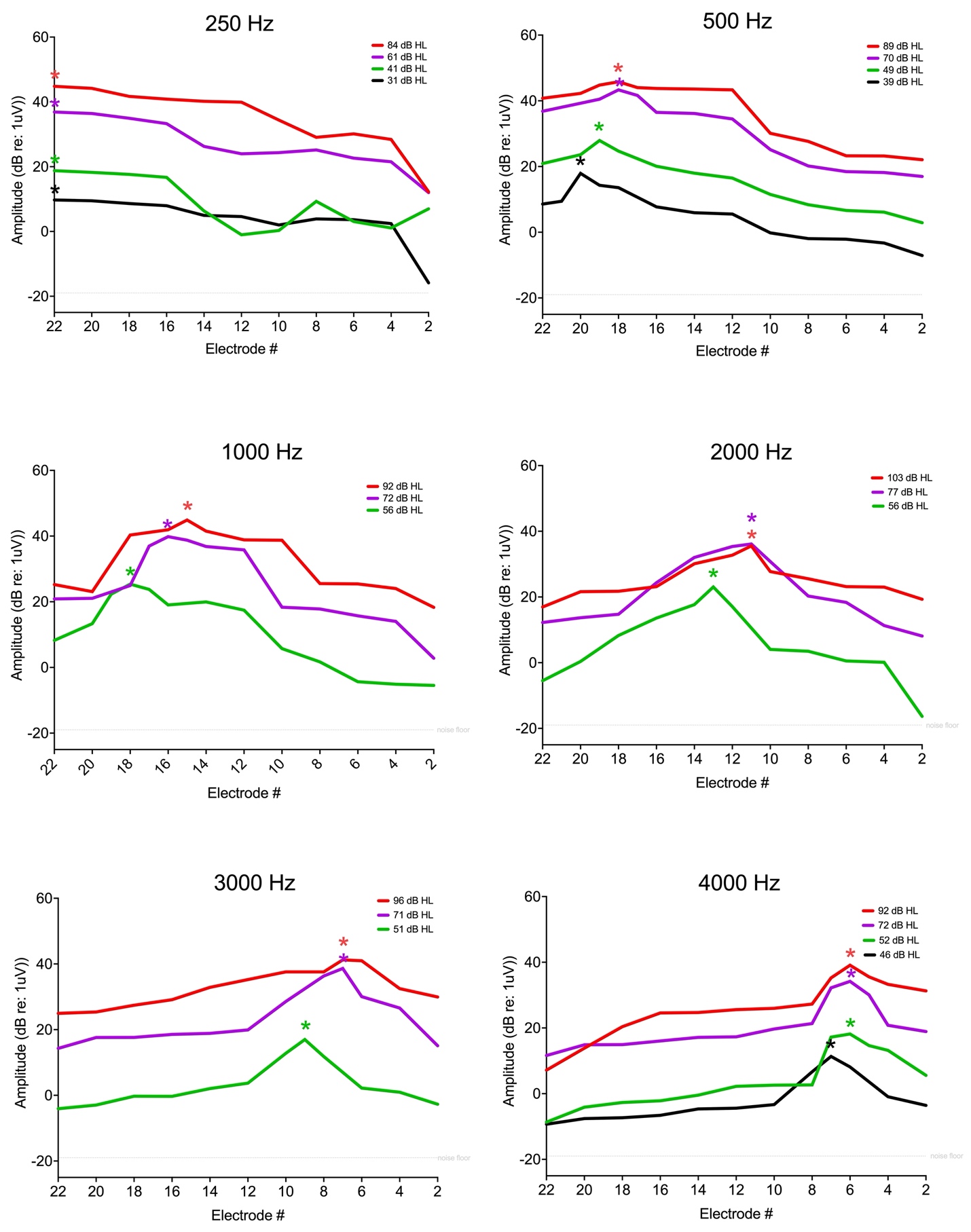

**Fig. S4. Influence of Intensity Stimulus Variation on Frequency-Position Mapping in Auditory Neuropathy Spectrum Disorder.** The tonotopic tuning derived from the intracochlear electrocochleography of a patient with auditory neuropathy spectrum disorder (characterized by significant preservation of cochlear hair cell function) was examined under varied stimulus intensities. The panels here illustrate apical shifts in frequency-position mapping at reduced stimulus levels for 500 Hz, 1000 Hz, 2000 Hz, 3000 Hz, and 4000 Hz frequencies. The depicted amplitudes correspond to the fast Fourier transformation amplitudes of the difference response, largely indicative of the cochlear microphonic tuning curve (outer hair cell tuning curve). Asterisks (*) denote the best frequency (BF) electrode for each frequency at a given stimulus intensity. Notably, response patterns at conversation-like stimulus levels mirrored those observed at peak stimulation levels, suggesting that frequency-position maps during conversation are more consistent with high-intensity, electrophysiologically-derived maps than those predicted by the Greenwood function.

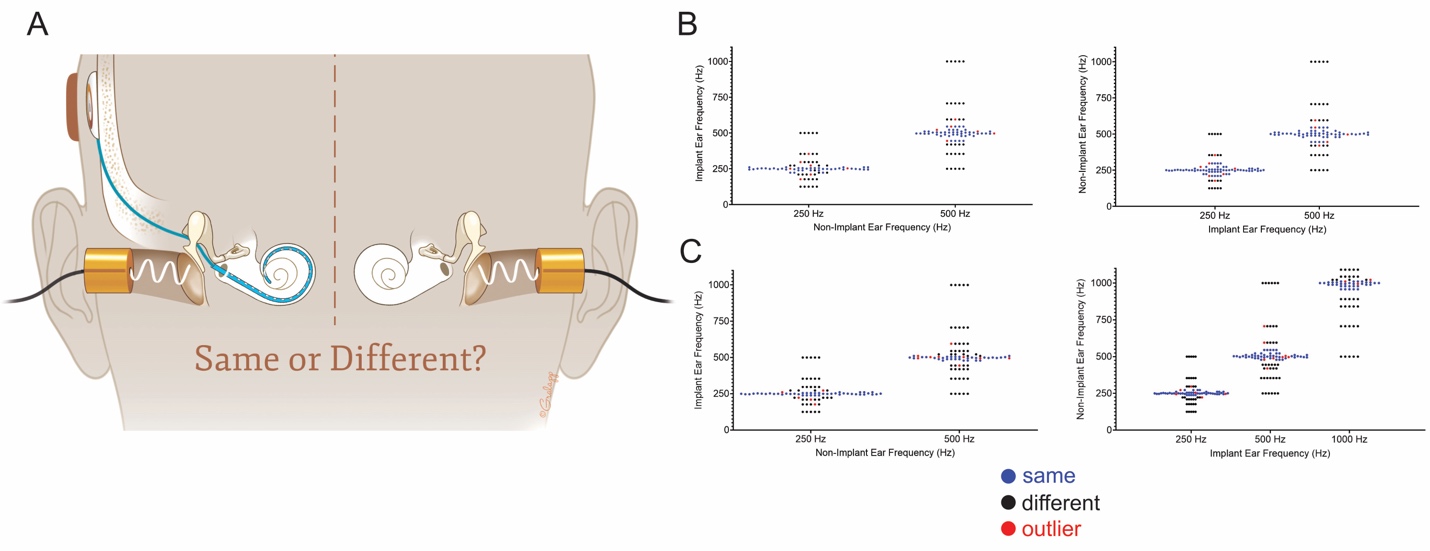

**Fig. S5. Pitch-discrimination testing to determine impact of presence of electrode on frequency-position map.** Pitch comparisons were obtained between acoustic stimuli presented sequentially to both the non-implanted ear and the implanted ear to determine whether the presence of the electrode had an impact on the acoustic-frequency position map. The cochlear implant processor was not used for this portion of the testing. (A) Two subjects were selected for this portion of the testing who had residual acoustic hearing preserved in the implanted ear and similar hearing in the contralateral, non-implanted ear. The acoustic stimuli were first balanced for loudness at all of the tested frequencies. Then one ear was held constant as the reference where a single, brief pure tone was delivered alternating randomly with varying frequency pure tones in the contralateral ear. The subject was then asked to indicate whether the pitches presented to each ear sequentially sounded the ‘same’ or ‘different’. In both subjects shown, the left graph in (B and C) shows the results where the non-implant ear was held constant and the acoustic stimulus was varied in the implant ear. The right graphs show the results where the implant ear was held constant and the non-implant ear was varied. The blue dots represent when the patient indicated that both pitches sound the same and the black dots represent when the patient indicated that both pitches sound different. The outliers in red were defined where the subject had one response that was different from the other 4 responses, which is commonly noted as a component of fatigue in pitch-discrimination testing. For subject 1 (B), there was a mean 1.30 semitone difference for the acoustic perception of pure tones for both ears. For subject 2 (C), there was mean 1.65 semitone difference for both ears. This testing indicates that the perimodiolar electrode did not impact the acoustic frequency tuning of the cochlea to a degree that could explain the shift shown between the electrophysiologically-derived frequency-position and those previously-established for the organ of Corti and spiral ganglion (2D).

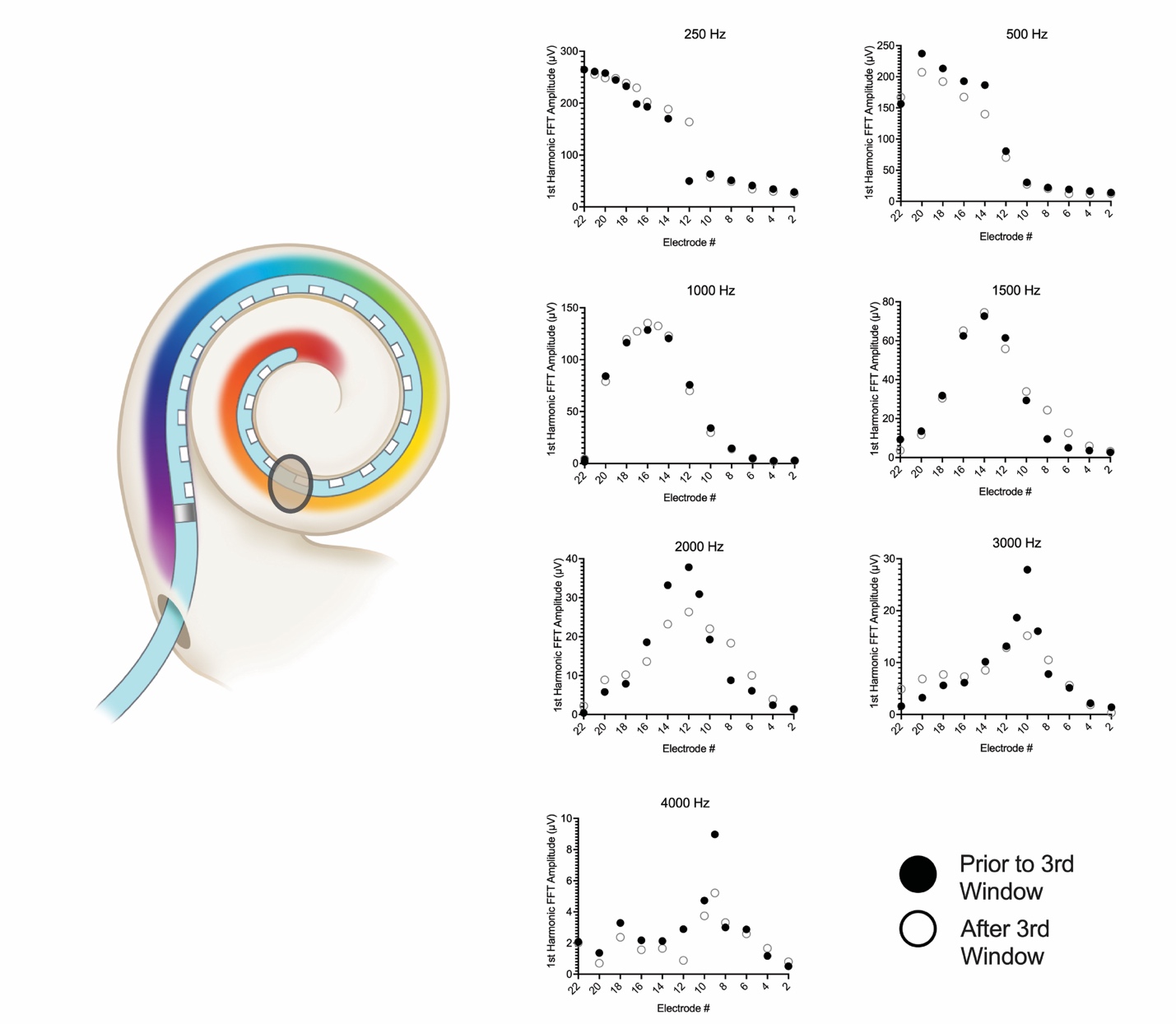

**Fig. S6. Third window fenestration and the frequency-position map.** Previous *ex vivo* experiments on tonotopy in humans have used a fenestration of the cochlear lumen to observe the traveling wave, but the potential impact of this artifact has not been studied. Impact of the third window on electrophysiologic recordings was evaluated in a human subject with excellent residual hearing who was undergoing a translabyrinthine craniotomy (which intentionally destroys residual hearing) for resection of a vestibular schwannoma. Prior to resection, the cochlea was approached identical to the electrophysiologic recordings performed previously and the best frequency (BF) was identified along the electrode array for 250-, 500-, 1000-, 2000-, 3000-, and 4000-Hz for the cochlear microphonic tuning curves (black dots in right panel). Following the recordings, a 1-mm diamond burr was used to create a fenestration near the upper cochlear turns (as shown in the left panel), and the CM tuning curves were generated for the individual frequencies (white dots in right panel).

**Supplementary Tables.**

**Table S1. Demographic, audiologic, and imaging information of fifty subjects tested to construct electrophysiologically-derived frequency-position map.**

|  | Mean ± STD or N (%) |
| --- | --- |
| Age (yrs) | 69.6 ± 18.3 |
| Gender | |
| Female | 31 (62.0) |
| Male | 19 (38.0) |
| Duration of hearing loss (yrs) | 29.0 ± 18.6 |
| Duration of severe-to-profound hearing loss (yrs) | 8.6 ± 10.4 |
| Etiology of hearing loss | |
| Unknown | 43 (86.0) |
| Sudden sensorineural hearing loss | 2 (4.0) |
| Congenital | 5 (10.0) |
| Low-frequency Pure Tone Average (LFPTA; 125, 250, 500 Hz; dB HL) | 56.3 ± 25.5 |
| Pure Tone Average (PTA; 500, 1000, 2000, 4000 Hz; dB HL) | 56.1 ± 20.8 |
| **CT 3-D Reconstructions** |  |
| Apical Electrode Insertion Angle (deg) | 404.3 ± 35.3 |
| Cochlear Diameter Center to Round Window (mm) | 9.2 ± 0.4 |

**Table S2. Comparative Analysis of Frequency-Position Functions: *In Vivo* vs. Greenwood Functions.**

| **Stimulus Frequency** | **Best Frequency Location (Degrees)** | **Average Greenwood Place (Degrees)** | **Greenwood Frequency (Hz)** | **Difference in Place (Degrees)** | **Difference in Frequency (Hz)** | **Octave difference** |
| --- | --- | --- | --- | --- | --- | --- |
| 500 | 325.6 ± 28.1 | 475.6 ± 23.2 | 1083.8 ± 221.4 | 150.0 ± 39.0 | 583.8 ± 221.4 | 1.1164 |
| 1000 | 211.3 ± 36.5 | 336.5 ± 14.1 | 2295.8 ± 625.0 | 125.2 ± 40.3 | 1295.8 ± 625.0 | 1.1899 |
| 2000 | 137.2 ± 28.0 | 224.7 ± 7.9 | 4007.5 ± 889.2 | 87.5 ± 28.7 | 2007.5 ± 889.2 | 1.0232 |
| 3000 | 115.0 ± 18.3 | 168.9 ± 3.8 | 4724.2 ± 746.3 | 53.9 ± 18.8 | 1724.2 ± 746.3 | 0.6207 |
| 4000 | 93.7 ± 16.6 | 133.1 ± 2.9 | 5666.4 ± 844.5 | 39.4 ± 17.3 | 1666.4 ± 844.5 | 0.5232 |
